# The WERA Cancer Center Matrix: Strategic Management of Patient Access to Precision Oncology in a Large and Mostly Rural Area of Germany

**DOI:** 10.1101/2024.03.10.24304028

**Authors:** Markus Krebs, Florian Haller, Silvia Spörl, Elena Gerhard-Hartmann, Kirsten Utpatel, Katja Maurus, Volker Kunzmann, Manik Chatterjee, Vivek Venkataramani, Imad Maatouk, Max Bittrich, Tatjana Einwag, Norbert Meidenbauer, Lars Tögel, Daniela Hirsch, Wolfgang Dietmaier, Felix Keil, Alexander Scheiter, Alexander Immel, Daniel Heudobler, Sabine Einhell, Ulrich Kaiser, Anja M. Sedlmeier, Julia Maurer, Gerhard Schenkirsch, Frank Jordan, Maximilian Schmutz, Sebastian Dintner, Andreas Rosenwald, Arndt Hartmann, Matthias Evert, Bruno Märkl, Ralf Bargou, Andreas Mackensen, Matthias W. Beckmann, Tobias Pukrop, Wolfgang Herr, Hermann Einsele, Martin Trepel, Maria-Elisabeth Goebeler, Rainer Claus, Alexander Kerscher, Florian Lüke

**Affiliations:** Comprehensive Cancer Center Mainfranken, University Hospital Würzburg, 97080 Würzburg, Germany; Department of Urology and Pediatric Urology, University Hospital Würzburg, 97080 Würzburg, Germany; Institute of Pathology, Friedrich-Alexander University Erlangen-Nuremberg, University Hospital Erlangen, 91054 Erlangen, Germany; Comprehensive Cancer Center Erlangen-EMN, 91054 Erlangen, Germany; Department of Medicine V, Hematology and Oncology, University Hospital Erlangen, 91054 Erlangen, Germany; Institute of Pathology, University of Würzburg, 97080 Würzburg, Germany; Institute of Pathology, University of Regensburg, 93053 Regensburg, Germany; Comprehensive Cancer Center Ostbayern, 93053 Regensburg, Germany; Department of Internal Medicine II, University Hospital Würzburg, 97080 Würzburg, Germany; Department of Internal Medicine III, Hematology and Oncology, University Hospital Regensburg, 93053 Regensburg, Germany; Comprehensive Cancer Center Augsburg, 86156 Augsburg, Germany; Department of Hematology and Clinical Oncology, Medical Faculty, University of Augsburg, 86156 Augsburg, Germany; Institute of Digital Medicine (IDM), Medical Faculty, University of Augsburg, 86156 Augsburg, Germany; Institute of Pathology and Molecular Diagnostics, Medical Faculty, University of Augsburg, 86156 Augsburg, Germany; Bavarian Cancer Research Center (BZKF), 91052 Erlangen, Germany; Department of Gynecology and Obstetrics, University Hospital Erlangen, 91054 Erlangen, Germany; Division of Personalized Tumor Therapy, Fraunhofer Institute for Toxicology and Experimental Medicine, 93053 Regensburg, Germany

**Keywords:** Cancer Care, Community Oncology, Molecular Tumor Board, Disparity in Healthcare, Healthcare Management

## Abstract

**Purpose:** Offering equal Patient Access to Precision Oncology (PO) is a major challenge of clinical oncologists and cancer center representatives. Here, we provide an easily transferable model adopted from strategic management science to assess the geographic impact of a cancer center – in terms of general cancer care and PO participation.

**Methods:** As members of the German WERA alliance, the cancer centers Würzburg, Erlangen, Regensburg and Augsburg merged care data regarding their geographical impact. Specifically, we examined the provenance of patients from WERA’s molecular tumor boards (MTBs) between 2020 and 2022 (n = 2243). As second dimension, we added the provenance of patients receiving general cancer care (termed Total Cancer Care, TCC) by WERA. Clustering our outreach along these two dimensions allowed us to set up a four-quadrant matrix consisting of postal code areas with referrals towards WERA. These areas were re-identified on a map of the Federal State of Bavaria and surrounding regions.

**Results:** In terms of positive MTB and general cancer care referrals, the WERA Matrix overlooked an active screening area of n = 821 postal code areas – representing about 50% of Bavaria’s spatial expansion and more than six million inhabitants. The WERA Matrix identified regions successfully connected to our outreach structures in terms of subsidiarity – with general cancer care mainly performed locally but PO performed in cooperation with WERA. At the same time, we detected postal code areas with a potential PO backlog – characterized by high levels of cancer care performed by WERA and low levels or no MTB representation.

**Conclusions:** The WERA Matrix provided a transparent portfolio of postal code areas, which helped assessing the geographical impact of our PO program. We believe that its intuitive principle can easily be transferred to other cancer centers.

## INTRODUCTION

Precision Oncology (PO) has demonstrated substantial clinical benefit for patients, especially in rare and hard-to-treat cancers [1–3]. Yet, providing equal access to PO is a major challenge for healthcare providers and government authorities. Several countries have established substantial infrastructure to foster Patient Access to PO – with a particular emphasis on patients living in rural areas. Japan and Norway for example have set up nationwide hospital networks with centralized molecular tumor boards (MTBs) [4–8]. For Germany, the “National Decade against Cancer” organized by the Federal Ministry of Education and Research aims to tackle obstacles in Patient Access to PO [9,10].

Nevertheless, not only healthcare policymakers are responsible for ensuring Patient Access – this job also falls within the purview of clinical oncologists. As part of the German NCT (National Center for Tumor Disease) network, the Comprehensive Cancer Centers of Würzburg, Erlangen, Regensburg and Augsburg have joined forces and established the WERA cancer center alliance. One of WERA’s central tasks is providing PO programs including clinical trials to its mainly rural catchment area, which covers the majority of the Federal State of Bavaria. In a first step, this task required capturing the geographical status quo of PO participation. As there is limited evidence on fostering PO in a community oncology setting [11,12], we chose an intuitive and hands-on approach. This initial analysis already revealed several “white spots” in our catchment area [13].

Here, we substantially extended our approach by adding general cancer care data from our four cancer centers. Merging both datasets allowed us to employ a four-quadrant matrix model from portfolio theory known as the Growth-Share Matrix [14]. Using the WERA Matrix enables our clinicians and cancer center representatives to better understand their regional PO impact and identify regions in our catchment area, which are currently not adequately covered.

## MATERIALS AND METHODS

### Data Sources

We merged postal code areas of the residences of WERA’s MTB patients between 2020 and 2022. Following harmonized standard operating procedures, our MTBs discuss patients diagnosed with an advanced cancer disease and no or limited treatment options left according to guideline recommendations [15,16]. Eligible for analysis were MTB patients with known postal code areas and residences in Germany. In addition, we merged postal code areas of the residences of all patients receiving cancer care at the WERA cancer centers. Using data from our four local cancer registries, we termed these referrals Total Cancer Care (TCC). In order to obtain a stable TCC catchment area and reduce the impact of potential outliers from single years, we calculated an average TCC across three years (ø 2018-2020). Table 1 summarizes the two datasets finally included and examined in our study.

**Table 1:**
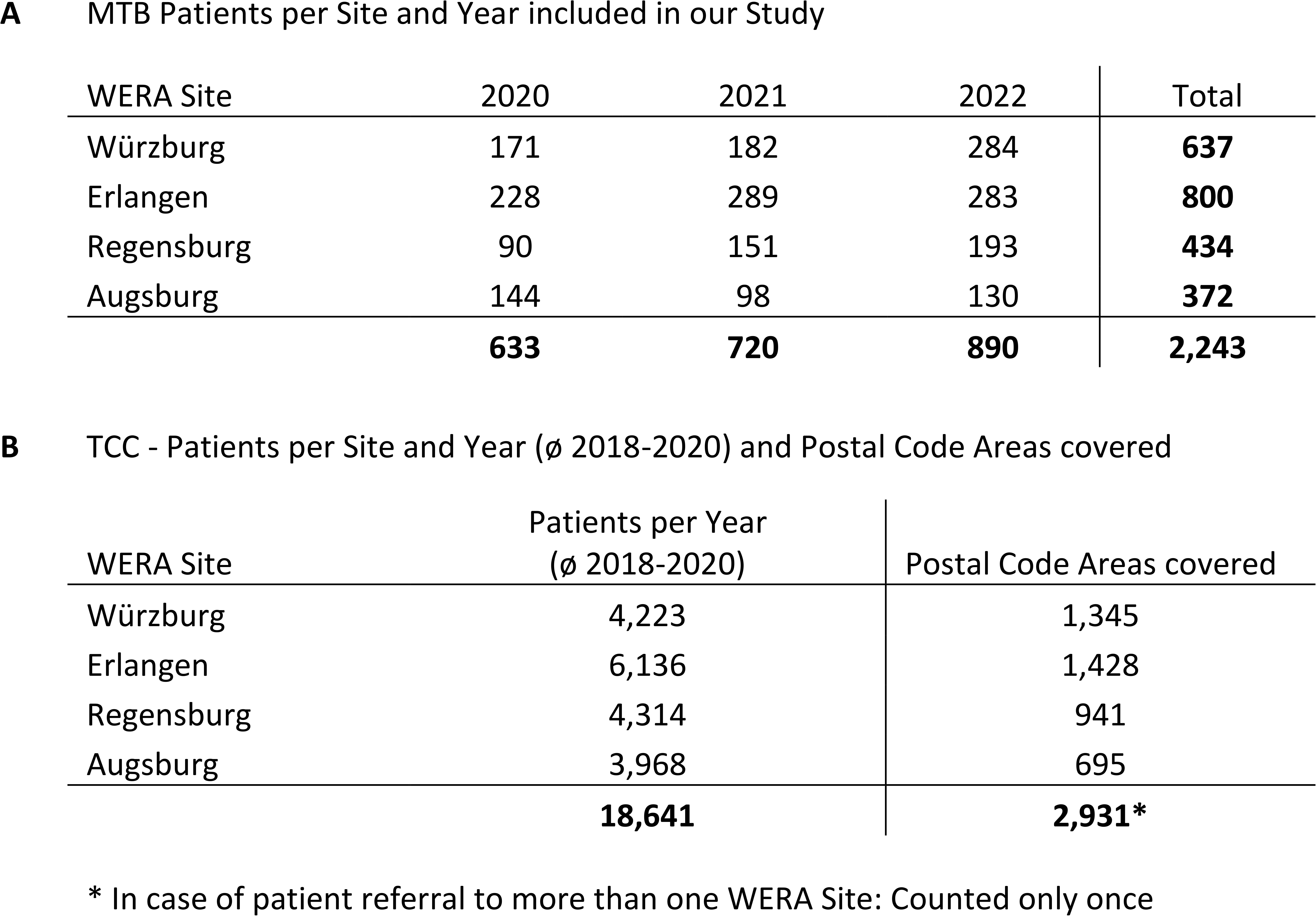
Cohorts included in this study. (A) Molecular tumor board (MTB) patients per WERA site and year analyzed. (B) Average number of patients receiving cancer care (termed Total Cancer Care, TCC) and number of postal code areas covered by each WERA site. TCC numbers were calculated as an average of the years 2018, 2019 and 2020.

Absolute MTB and TCC patient numbers per postal code area were divided by local population numbers – resulting in MTB and TCC patient numbers per 100,000 inhabitants and postal code area. Population densities per postal code area were collected from a freely accessible source (htps://www.suche-postleitzahl.org/downloads). This database combines information from German statistical offices as part of the “Zensus 2011” initiative (https://www.zensus2011.de) with geospatial information from OpenStreetMap (https://www.openstreetmap.org).

Due to the retrospective nature and the exclusive utilization of anonymized data, analysis of MTB and TCC patients was in accordance with German General Data Protection Regulation (GDPR) and legislation, specifically the Bavarian Hospital Act (“Bayerisches Krankenhausgesetz”).

### Formal Analysis and Graphical Illustration

We merged and analyzed the residences of MTB and TCC patients by using Microsoft® Access® 2016 (version 16.0.5224.1000, Redmond, WA, USA). For visualization of geographical information, we employed QGIS, an open-source information system (QGIS Development Team; under license of GNU General Public License, Version 3.26.3). Stepwise color-coding of maps within QGIS employed the Jenks optimization method, which aims to minimize variance within groups while maximizing variance between groups [17]. The WERA Matrix was visualized using Microsoft® PowerBI® (version 2.118.621.0 32-bit (June 2023), Redmond, WA, USA).

### Portfolio Analysis and Strategic Assessment of Outreach Measures

For our study, we compared two dimensions: the regional distribution of MTB patients and the regional distribution of TCC patients from the WERA cancer center consortium. The latter reflects established and comparably stable streams of cancer patients towards our four centers.

Originally, the Growth-Share Matrix (Figure 1A) helped enterprises to visualize and manage their product portfolio – by combining the market share of a certain product with the dynamics (growth perspective) of its market environment [14]. While the x-axis reflects the current strength of a product in a given market, the y-axis reflects its innovative capability. Adopting and modifying this management tool from a portfolio of consumer goods to a portfolio of postal code areas (Figure 1B) allowed us to combine WERA’s share in total cancer care (x axis) with the local representation of MTB patients (y axis).

**Figure 1:**
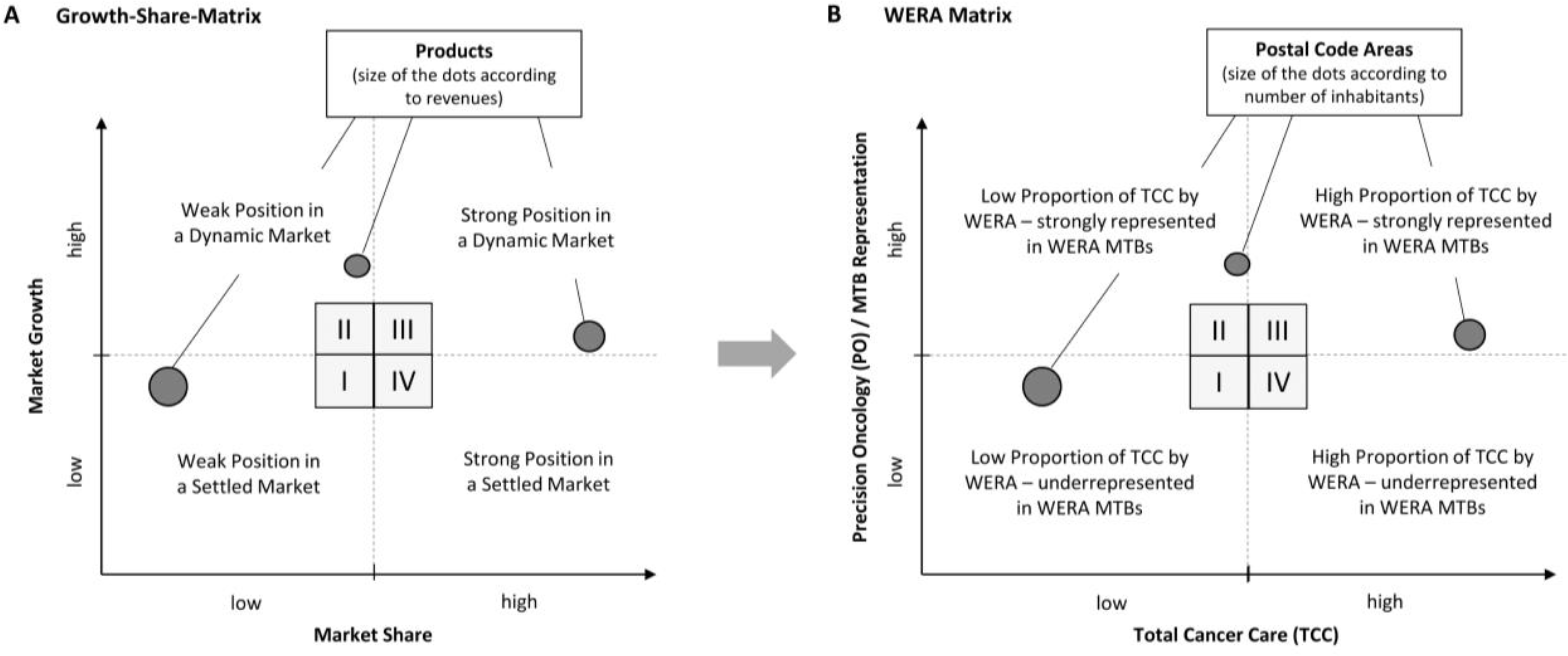
The WERA Matrix derived and modified from Strategic Portfolio Management. (A) Growth-Share Matrix depicting market shares and market growth rates for specific products. (B) Merging WERA’s proportion in Total Cancer Care (TCC, x-axis) with the local MTB (molecular tumor board) representation (y-axis) for each outreach postal code area.

## RESULTS

### Longitudinal Screening of Patient Access to WERA MTBs

Building upon our earlier analysis [13], we initially screened the geographical development of our MTB outreach by mapping relative MTB representation (patients per 100,000 inhabitants) per postal code area. While the numbers from the years 2020 to 2022 still represented a limited database, the resulting map (Figure 2) already provided insights into the longitudinal evolution of our PO program. Regarding “white spots” from the years 2020 and 2021, some regions newly emerged as MTB referrals in 2022 (indicated by bright blue frames around postal codes areas) – thereby indicating an organic growth of our program. For example, regions close to Ingolstadt (Figure 2, *) and Straubing (Figure 2, §) were newly established. At the same time, some areas not covered within our previous analysis still did not appear in our updated analysis – such as regions close to the Czech Border on the eastern rim of our catchment area (Figure 2, #).

**Figure 2:**
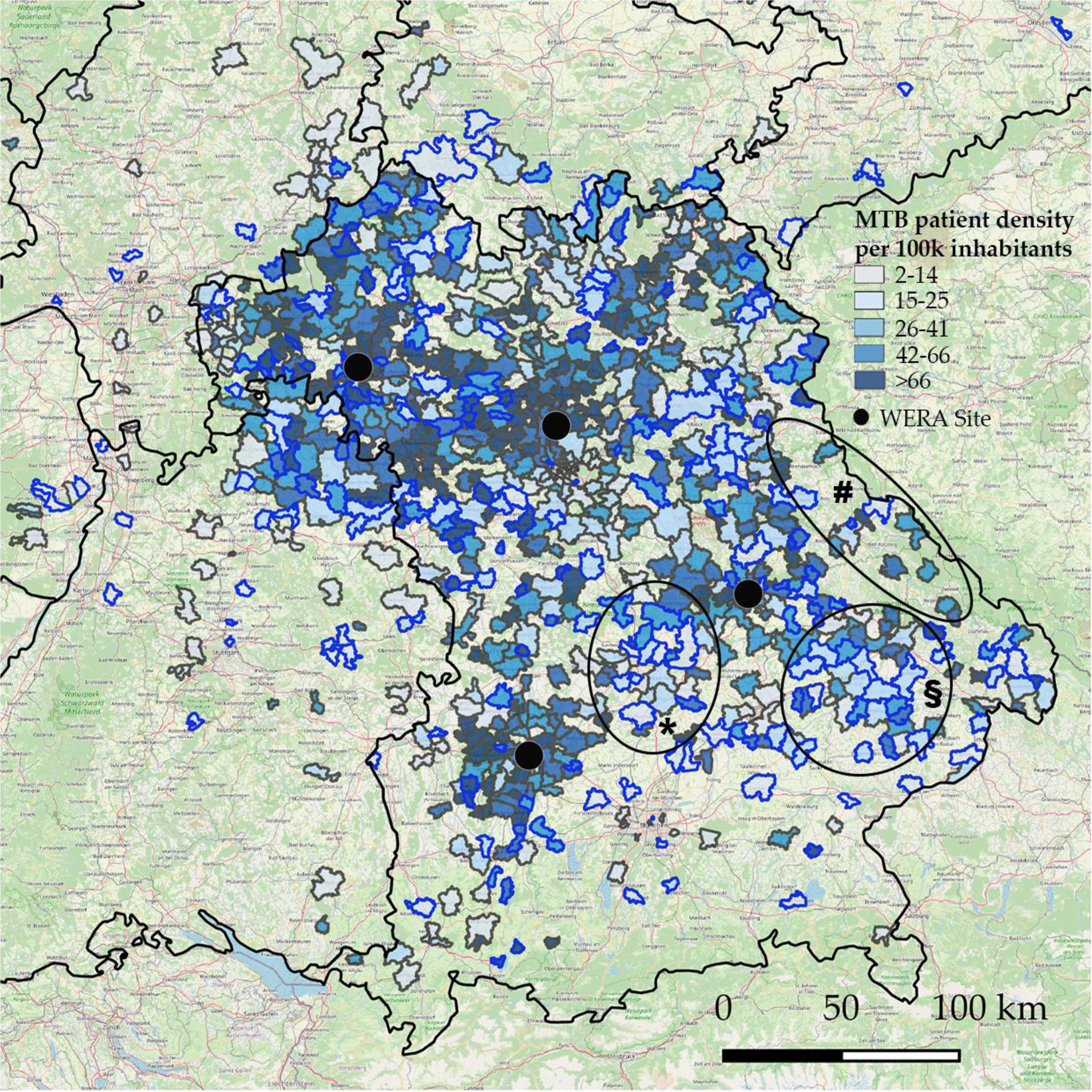
The WERA MAP – Federal State of Bavaria with neighboring regions and color-coded development of MTB (molecular tumor board) representation per 100,000 inhabitants and postal code area. Bright blue frames around postal code areas indicate first appearance in the year 2022 – such as regions close to Ingolstadt (*) and Straubing (§). # indicates a weakly covered part of eastern Bavaria close to the Czech border (“Bayerischer Wald”).

Subsequently, we explored the extent to which patients from these remaining “white spots” availed cancer care services from WERA, irrespective of tumor type, clinical stage, and treatment modalities. To achieve this, we merged cancer care (TCC) data sourced from our four cancer registries and then visually represented the relative prevalence of TCC patients (per 100,000 inhabitants and postal code area) on a map encompassing the Federal State of Bavaria and its adjoining regions. By empirically establishing a visualization threshold of 15 TCC patients per 100,000 inhabitants, postal code area, and year, a coherent catchment area was delineated (Figure 3). Therefore, our analysis focused on the primary region where WERA provides cancer care. With the exception of Upper Bavaria, the region surrounding Munich and the southeastern area proximate to the Alps (Figure 3, *), WERA’s catchment area encompassed a significant portion of Bavaria and certain neighboring regions—specifically, the northern part of Baden-Württemberg (Figure 3, §).

**Figure 3:**
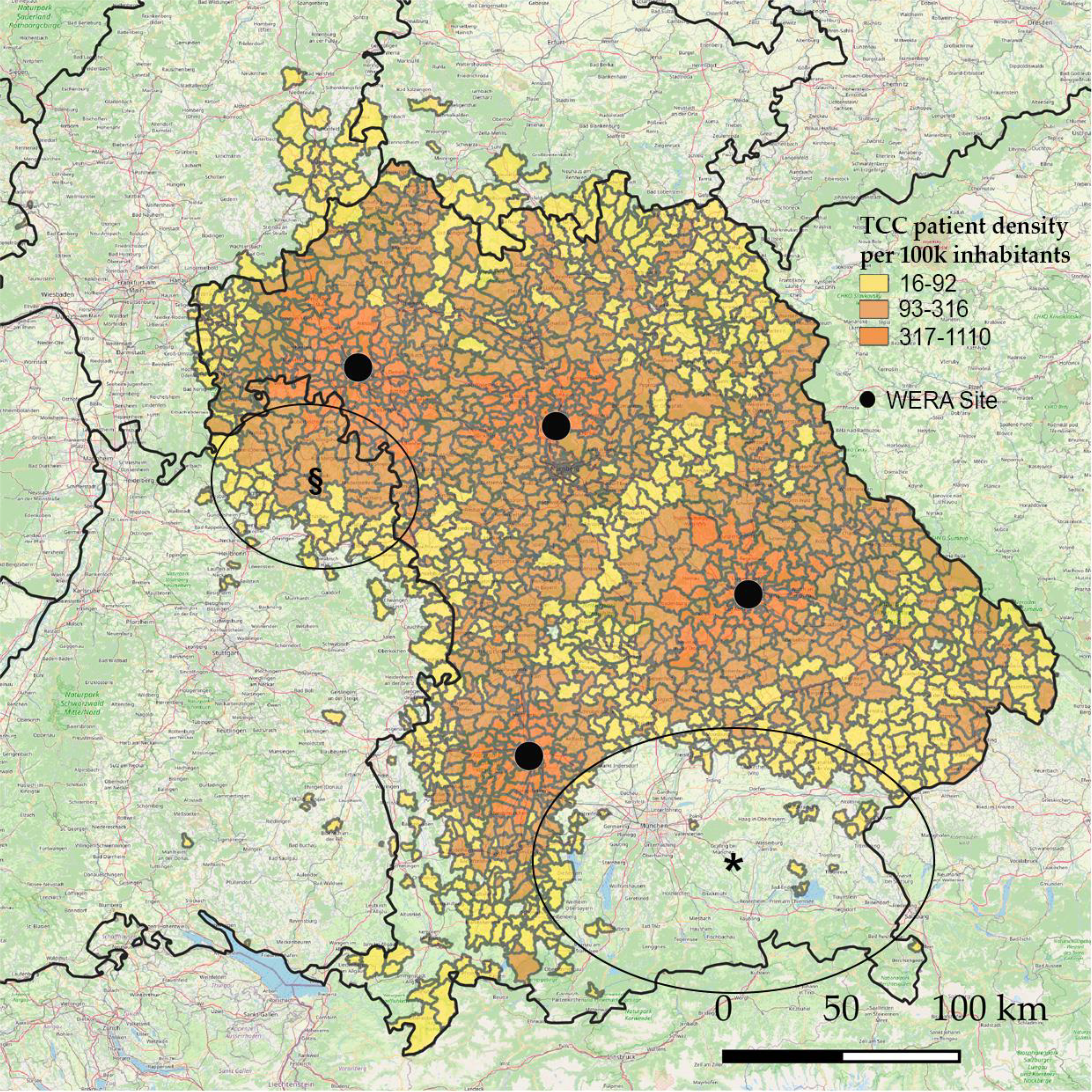
Total cancer care (TCC) performed by the WERA cancer centers – Regional visualization (per postal code area) of cancer care patients weighted with local population densities; patient density is depicted as patients treated at a WERA center per 100,000 inhabitants and year. § indicates the northern part of the federal state of Baden-Württemberg. * indicates Upper Bavaria including the region around Munich (“Oberbayern”).

When comparing MTB representation and TCC representation on the level of postal code areas, we detected substantial discrepancies between both variables. For example, the eastern rim (Figure 2, #), which was weakly covered in terms of MTB representation, was well represented in terms of TCC (Figure 3). These findings led us to establish a broader graphical approach for all postal code areas belonging to our pre-defined catchment area. As outlined in Figure 1, we adopted and modified the Growth-Share Matrix from strategic portfolio management [14] for this approach.

### Illustrating Regional Imbalances in MTB and TCC representation: the WERA Matrix

For the WERA Matrix (Figure 4), we plotted postal code areas (size of the dots depending on population size) along the two dimensions TCC representation (per 100,000 inhabitants) and MTB representation (per 100,000 inhabitants). Dots on the x-axis (Figure 4, y = 0) represented n = 920 postal code areas with TCC > 15 patients per 100k inhabitants and year but no MTB referral. In contrast, dots on the y-axis (Figure 4, x = 0) described n=128 postal code areas with MTB referrals but no TCC referrals – which mostly represented MTB referrals from beyond our catchment area. Our graphical approach included n = 821 postal code areas with simultaneous TCC and MTB referrals – overall representing a screening area of 34,458.7 square kilometers (about 50% of the Federal State of Bavaria) and 6,367,915 inhabitants.

**Figure 4:**
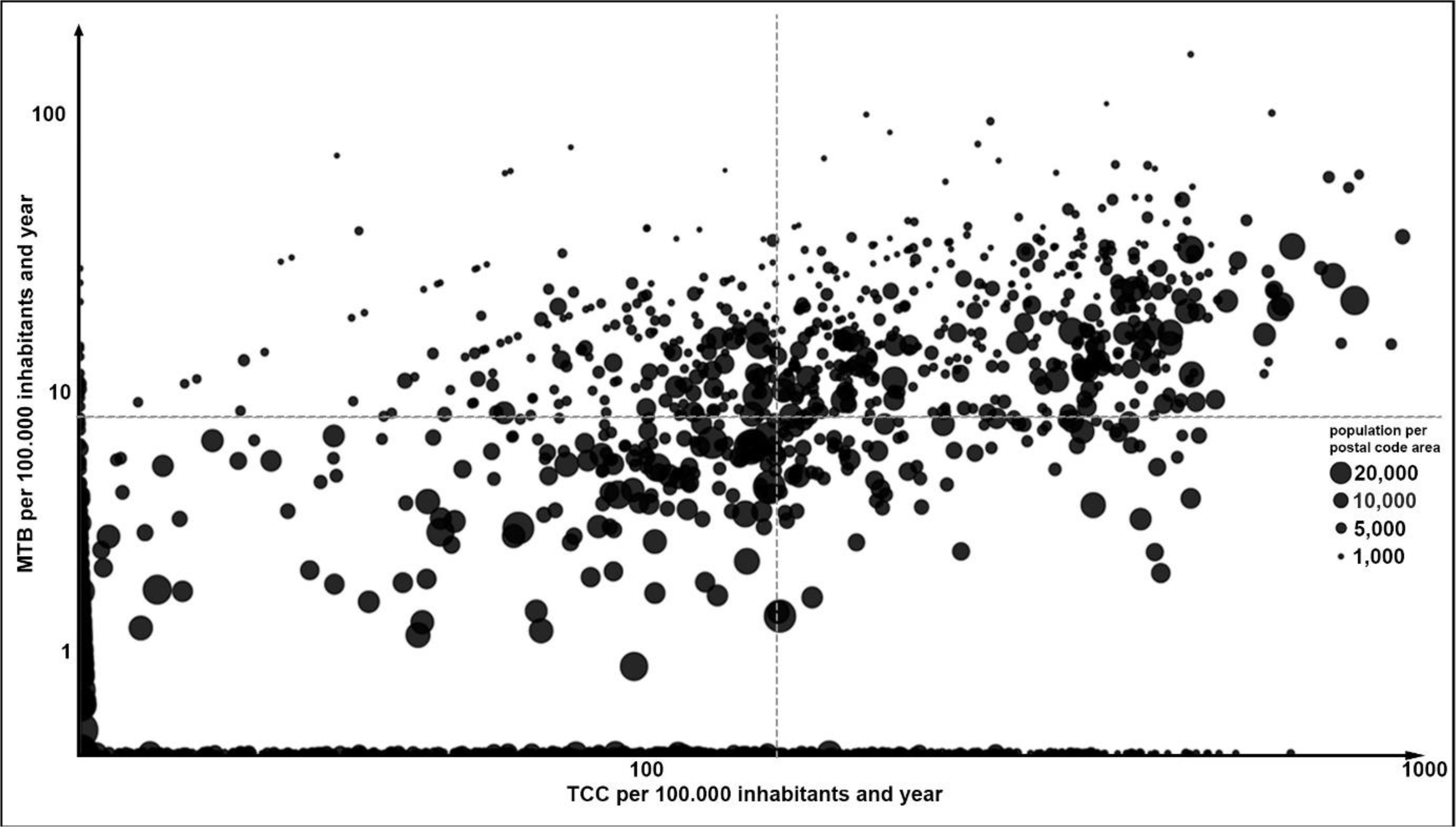
The WERA Matrix - dot plot of postal code areas with patient referral towards the WERA cancer centers along the two dimensions Total Cancer Care (TCC) and molecular tumor board (MTB) representation (per 100,000 inhabitants and year). Each dot describes one postal code area – with dot sizes depending on local population. TCC and MTB representation are depicted on a logarithmic scale. Dashed grey lines describe median TCC and MTB representation for both datasets.

As we aimed to assess MTB representation in WERA’s TCC catchment area, we excluded postal code areas with TCC referrals lower than 15/100.000 inhabitants. The remaining postal code areas of the WERA Matrix belonged to one of four quadrants – with median TCC representation and median MTB representation serving as cut-off values. Moreover, this semi-quantitative approach allowed us to re-identify the postal code areas of each quadrant on a map of Bavaria and its surrounding regions (Figure 5).

**Figure 5:**
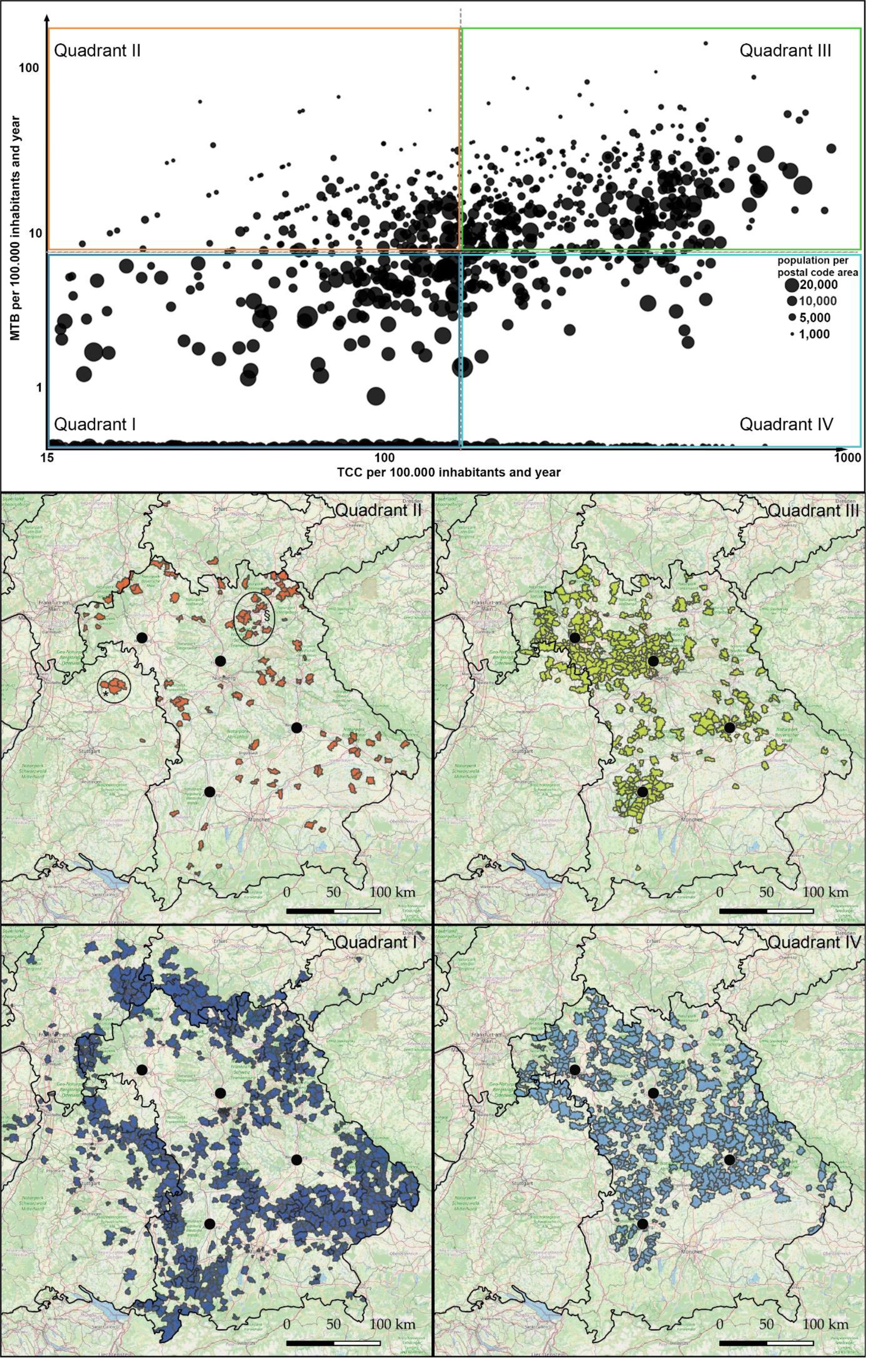
The WERA Matrix – assignment of postal code areas to one of four quadrants and geographical re-identification on a map of the Federal State of Bavaria with surrounding regions. Quadrant I: TCC low (local TCC representation < median TCC representation) and MTB low (local MTB representation < median MTB representation). Quadrant II: TCC low (local TCC representation < median TCC representation) and MTB high (local MTB representation > median MTB representation). Quadrant III: TCC high (local TCC representation > median TCC representation) and MTB high (local MTB representation > median MTB representation). Quadrant IV: TCC high (local TCC representation > median TCC representation) and MTB low (local MTB representation < median MTB representation). * describes quadrant II areas close to Bad Mergentheim. § describes quadrant II areas close to Bayreuth.

Areas characterized by relatively low TCC and low MTB representation (lower left quadrant I, depicted in dark blue) mainly were situated in the periphery of our catchment area. In addition, some of these areas lay halfway between WERA sites. Quadrant II postal code areas (upper left, depicted in orange) were characterized by a relatively low TCC combined with a high MTB representation. This quadrant contained regions and cities like Bad Mergentheim (Fig 5 *) and Bayreuth (Figure 5 §), which already are close cooperation partners within our cancer center network. In contrast, postal codes areas belonging to quadrant III (upper right, depicted in green) usually were located close to our four WERA cancer centers. Quadrant IV (lower right, depicted in light blue) finally contained postal code areas characterized by a strong TCC representation along with a relatively weak representation in WERA MTBs. Several of these regions were located in the periphery of our catchment area. However, some quadrant IV areas also emerged in short distance to our cancer centers – indicating potential gaps in terms of PO coverage close to our cancer centers.

Regarding the overall distribution of postal code areas in the WERA Matrix (Figure 5), many of them clustered close to the center. This central clustering potentially limits the validity of our graphical approach which exclusively assigns regions towards only four quadrants. Therefore, we aimed to improve the granularity of our approach by introducing customized gates to identify sub-clusters within quadrants (Figure 6). For these bona fide gates, we applied the following coordinates (in x / 100.000 inhabitants and year):

**Figure 6:**
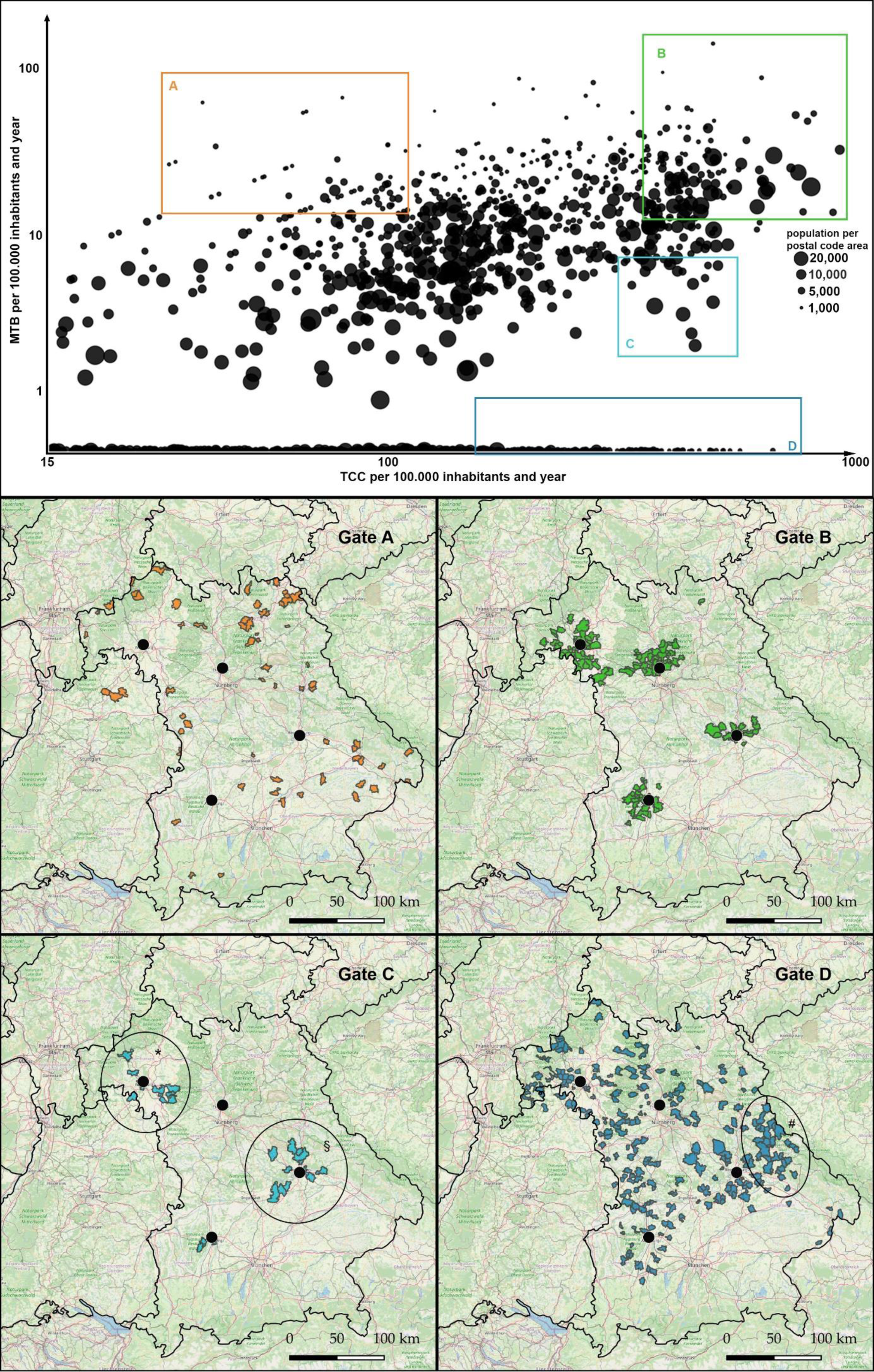
The WERA Matrix – assignment of postal code areas to predefined Gates / sub-clusters and geographical re-identification on a map of the Federal State of Bavaria with surrounding regions. Quadrant C specifically contained areas close to Würzburg (*) and Regensburg (§). The eastern rim of our catchment area (“Bayerischer Wald”, #) was particularly represented within gate D.

- Gate A (Orange): 27 < TCC < 104; 13 < MTB < 80
- Gate B (Green): 379 < TCC < 1200; 12 < MTB < 160
- Gate C (Light Blue): 315 < TCC < 600; 2< MTB < 8,5
- Gate D (Dark Blue): 220 < TCC < 1000; 0 < MTB < 1

As already mentioned for quadrant II, postal code areas belonging to Gate A represented regions close to cooperating outreach partners. As a sub-cluster of quadrant III, Gate B mainly re-identified urban regions very close to the WERA hubs – with a strong representation in terms of both TCC and MTB participation. Regarding “white spots” and imbalances in Patient Access, the two remaining gates C and D appear most interesting. Of note, gate D represented the clearest discrepancy – with regions characterized by strong TCC representation while having no patients discussed in WERA’s MTBs. After geographical re-identification, these specific regions were not randomly distributed across our catchment area. Instead, some of these regions such as the eastern rim (Figure 6 #, “Bayerischer Wald”) also clustered geographically, implying that regional socioeconomic determinants could contribute to the imbalance between TCC and MTB representation. However, “white spots” were not only located in rural areas far away from WERA’s cancer centers – instead, we detected them in urban regions and suburbs of Würzburg (Figure 6 *) and Regensburg (Figure 6 §).

## DISCUSSION

### Longitudinal Assessment of Geographical PO Coverage

We previously defined the joint PO coverage area of our cancer centers by merging patient care data from our four MTBs [13]. Thus, we showed successful outreach structures but also regional “white spots” in terms of MTB representation. In this study, we assessed the further geographical development of our PO coverage by adding MTB patient data from 2022. Upon closer examination of former “white spots” [13], we discovered that some of them were filled recently, particularly in regions close to the cities of Ingolstadt and Straubing – thereby reflecting successful outreach efforts and, in general, the growth of our PO program. In contrast, some “white spots” such as the eastern rim of our catchment area (“Bayerischer Wald”) maintained to be uncovered. To find out whether filling these gaps is merely a question of organic growth and thus a question of time or whether there are systemic obstacles to Patient Access, we needed to expand and modify our previous approach. For this, we added regional information on general cancer care (TCC) performed by WERA cancer centers.

### The WERA Matrix as a Strategic Outreach Management Tool

For merging PO coverage and TCC coverage, we adopted the Growth-Share Matrix from strategic portfolio planning [14] in order to assess the development of local MTB representation not only regarding local population, but also regarding established patient streams receiving cancer care in the WERA network. The resulting WERA Matrix attributed postal code areas to one of four quadrants – each representing a certain real-life scenario (Figure 7).

**Figure 7:**
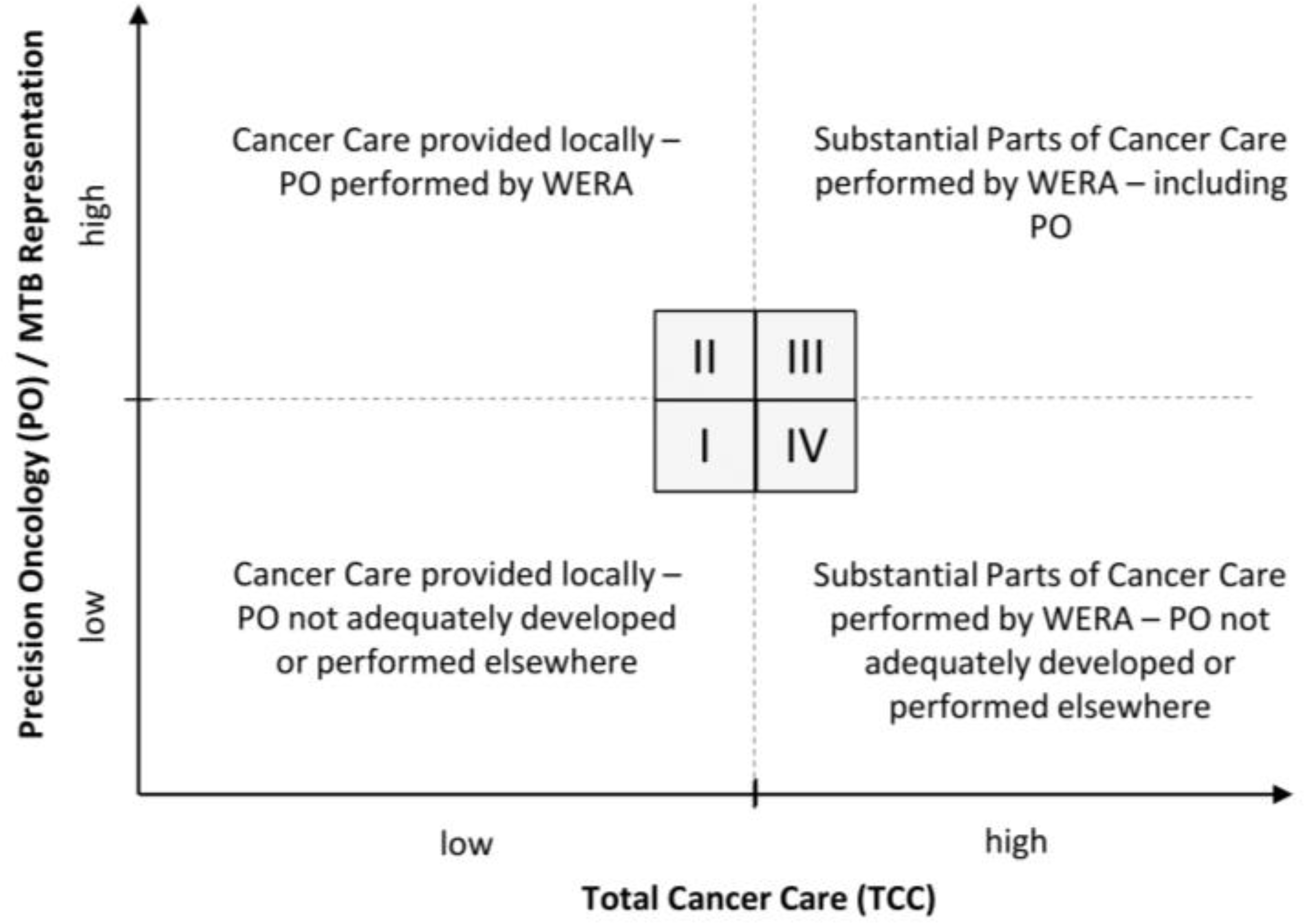
Potential real-life scenario for each quadrant of the WERA Matrix.

Geographical re-identification and visualization of postal code areas belonging to each quadrant allowed us to draw conclusions about local determinants of Patient Access. For example, postal code areas belonging to quadrant II could serve as an example for successful and efficient outreach structures in community oncology – with general cancer care mainly performed locally but PO performed in cooperation between local care providers and the WERA cancer centers – reflecting functioning PO outreach structures. In contrast, postal code areas belonging to quadrant IV appear promising for future outreach activities – as this quadrant contains regions with strong and established patient referral towards WERA centers in terms of TCC, while being underrepresented in terms of PO (MTB) representation. Part of the explanation for this discrepancy may lie in insufficient information and awareness among local physicians and patients. Moreover, technical and organizational obstacles could be responsible for this discrepancy.

To add further granularity to our semi-quantitative matrix approach, we selected clusters of postal code areas within a given quadrant. Thus, we identified “urgent white spots” – regions with a strong referral of patients towards our centers for TCC but no MTB representation at all. Most importantly, these areas were not exclusively located far away from our centers. Instead, we also detected them in close vicinity to our cancer centers. Moreover, these “urgent white spots” were not evenly distributed across our catchment area but clustered in specific areas, implying that common socioeconomic determinants could prevent patients from accessing suitable PO measures. In the future, we have to examine each of these “white spots” in order to understand individual local factors contributing to imbalances between MTB and TCC representation. Ultimately, this deeper understanding of local obstacles to Patient Access will help us developing tailored and effective outreach measures.

### Strengths and Limitations of our Approach

Our study has several limitations. For example, established streams of cancer patients do not necessarily reflect a need for PO – as some cancers currently have a limited spectrum of druggable targets. Moreover, certain subgroups such as patients with a localized disease, or patients in palliative care might not benefit from PO measures. Yet, we did not intend to use TCC as a surrogate for immediate PO need. Instead, it reflects the willingness of local patients – and local physicians – to receive cancer care from WERA cancer centers.

From a methodological perspective, portfolio analysis using the Growth-Share Matrix [14] has had a huge impact on strategic management especially in the last century. Yet, later research has questioned its applicability in a business setting partly due to inherent over-simplification [18]. At the same time, portfolio management techniques, including the Growth-Share Matrix, are still used in many companies, especially when it comes to setting strategic goals for business units and visualizing the strategic status quo [19]. In this light, we are convinced that a certain degree of simplification and catchiness is required to introduce a management tool as a novel frame of thinking in our clinical setting. Finally, we wanted to shed light on the fact that delivering Patient Access to PO also represents a considerable management task for clinical oncologists and representatives of cancer centers [11,20,21] – which should be supported by transparent and easily accessible management tools.

## Data Availability

All data produced in the present study are available upon reasonable request to the authors.

